# Headache-associated photophobia is more prevalent during winter: a cross-sectional study from a pediatric headache clinic

**DOI:** 10.1101/2025.10.14.25338019

**Authors:** Carlyn Patterson Gentile, Geoffrey K. Aguirre, Andrew D. Hershey, Christina L. Szperka

## Abstract

**Aim:** Sensitivity to light and noise is commonly associated with headache. It is unclear if exposure to more intense sensory environments, or exposure to more restricted sensory environments leading to intolerance, worsens sensory sensitivity more over time. We aimed to determine if photophobia is more common during the winter months when daily light exposure is reduced.

**Methods:** We conducted a single-center cross-sectional study from a pediatric headache clinic in the Philadelphia area assessing patient-reported headache-related photophobia and phonophobia from an outpatient headache intake questionnaire. Rates were compared in the 3 months surrounding the winter and summer solstice, which has an average daylight difference of 5 hours between these time windows. Monthly symptom rates were fitted to a sinusoid across the 12-month calendar to characterize seasonal variation.

**Results:** A total of 2,040 headache intake questionnaires were included in the analysis. Headache-associated photophobia was significantly more common in the winter compared to summer (OR 1.38, p = 0.018), but this seasonal variation was not observed for phonophobia (p = 0.698).

**Conclusion:** Headache-related photophobia is more prevalent near the winter, compared to the summer, solstice.

## Introduction

Sensory sensitivity, including photophobia and phonophobia, is commonly associated with headache and can be seen in migraine, tension-type headache, and post-traumatic headache, among others.^1^ One may predict that exposure to brighter light and louder noise would significantly worsen sensory sensitivity and should be avoided. However, it has also been suggested that reduced exposure to light or noise over time may lead to worsening intolerance. This has clinical implications, whether avoidance or graded exposure should be the main strategy to manage sensory sensitivity.^2^ In temperate climates, individuals are exposed to significantly more light in the summer compared to the winter.^3,4^ This difference offers an opportunity to perform a natural study to assess if headache-related photophobia is more likely to be reported in the summer when light exposure is the highest, the winter when light exposure is reduced, or shows no seasonal variation. If photophobia does indeed have a seasonal variation due to differences in daily light exposure, phonophobia should not be affected because there is not a comparable variation in noise exposure. As associated symptoms evolve across development with rates of vomiting decreasing, and rates of photophobia and phonophobia increasing with a notable change around age 12,^5^ we chose to focus on older youth and young adults to remove this potential source of variability.

We determined rates of photophobia and phonophobia reported by youth and young adults (12 years and older) on a headache intake questionnaire from general neurology and headache clinics. We compared rates in the 3 months surrounding the winter solstice (November – January) when the average length of daylight is 9.7 hours, to the summer solstice (May – July) when the average daylight length is 14.7 hours.^6^ We hypothesized that photophobia would be more prevalent closer to the winter solstice compared to the summer solstice when light exposure is lower, while phonophobia would not show seasonal variation.

## Methods

This was a single-center cross-sectional study based on standardized patient-reported headache intake questionnaire data from the Children’s Hospital of Philadelphia (CHOP) headache registry.^7^ Patient survey data were managed using REDCap electronic data^8,9^ capture tools hosted at CHOP. The institution’s Institutional Review Board approved the extraction of the data from the electronic health record into a research registry, with a waiver of consent and assent to maximize generalizability. Sample size was based on data availability, and we have found sample size was sufficient based on past studies using the headache registry.

Intake questionnaires were included if they were filled out between November 2022 and August 2024 and patients were 12 years and older. The inclusion time window was chosen as completion rates improved significantly in November 2022 onward due to quality improvement measures.^7^ Given that a low rate of forms were not fully completed (4.9%), no adjustments for missing data were made. Data were collected on the presence or absence on headache-associated photophobia and phonophobia.

Data analysis was performed in Matlab (Mathworks, Natick, MA) using publicly available custom software. Rates of photophobia and phonophobia were reported as proportion of patients and are graphically represented with 95% confidence intervals. Univariable binomial regression analysis was used to determine the effect of winter versus summer solstice (predictor) on the presence versus absence of photophobia or phonophobia (outcome). Significance was defined as p < 0.05. To further characterize seasonal variation, monthly rates of photophobia and phonophobia were fitted with a sinusoidal function with a period of 12 months and R squared was calculated for goodness-of-fit.

## Results

Of the 3,207 headache intake questionnaires started between November 2022 and August 2024, 2,040 (63.6%) were from patients 12 years and older that were used in the analysis. Patients were 72.5% female and were a median age of 15 [IQR 13, 16].

Of all questionnaires included in the analysis, 1,044 were collected either between November – January (57.1%) or May – July (42.9%), which were used in the comparison between the winter and summer solstice. Rates of photophobia were 73.4% in the winter, and 66.7% in the summer, while rates of photophobia (56.4% vs. 55.1%) did not show as large of a discrepancy (Figure 1a). Univariable binomial regression analysis was used to determine if rates of photophobia and phonophobia differed based on proximity to the winter versus summer solstice. Around the winter solstice, respondents were significantly more likely to report headache-associated photophobia (OR 1.38 [95%CI 1.06, 1.81], p = 0.018). This was not observed for phonophobia (OR 1.05 [95%CI 0.82, 1.35], p = 0.698). We then fitted the monthly rates of photophobia and phonophobia with a sinusoidal function (photophobia is shown in Figure 1b).

**Figure 1:**
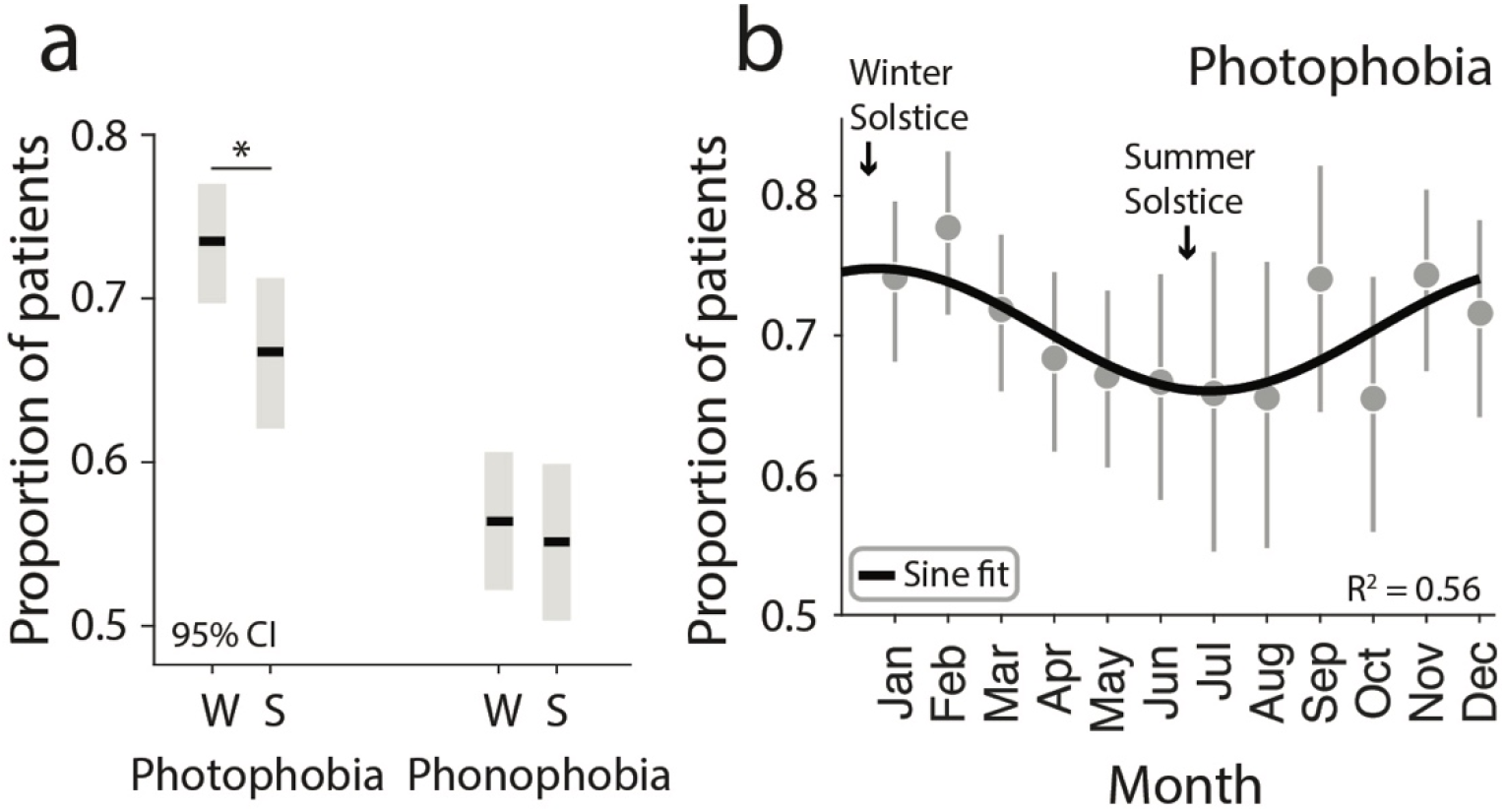
Prevalence of photophobia and phonophobia by season. (a) The rates of photophobia and phonophobia over the 3 months surrounding the winter solstice and summer solstice is shown with 95% confidence intervals. (b) Monthly rates of photophobia fitted to a sinusoidal function. * indicates p < 0.05. W = winter solstice, S = summer solstice, Jan = January, Feb = February, Mar = March, Apr = April, Jun = June, Jul = July,Aug = August, Sep = September, Oct = October, Nov = November, Dec = December.

The sinusoidal function was a moderately good fit for photophobia across the year suggestive of seasonal variation (R^2^ = 0.56), but a poor fit for phonophobia (R^2^ = 0.18).

We considered that differences in the prevalence of photophobia between the winter and summer solstice could be explained by being in versus out of school. However, rates of photophobia started decreasing in March during the school year and steadily declined reaching their nadir at the Summer Solstice, making this explanation unlikely.

## Discussion

We found that rates of reported headache-related photophobia in a pediatric headache intake questionnaire showed seasonal variation, with greater rates observed near the winter solstice. This finding demonstrates that higher rates of photophobia are reported at the time-of-year associated with a decrease in daily light exposure, which suggests that exposure to lower light levels reduces light tolerance. The prevalence of headache and migraine symptoms tend to increase in temperate climates during the fall and winter,^10–12^ which could explain an increase in photophobia during the winter. However, phonophobia did not demonstrate seasonal variation, making this explanation less likely. Our study is contrary to findings from the sub-arctic, where rates of photophobia and migraine are higher in the bright summer months,^13^ which may reflect differences in extreme versus moderate fluctuations in daylight length.

The seasonal variation we observed in photophobia could not be fully accounted for by differences in being in versus out of school. Rates of photophobia started decreasing in March during the school year and steadily declined reaching their nadir in June, which is the start of summer break. We did see an increase in photophobia in the month of September at the start of the school year. We speculate that this may result from a relative decrease in light exposure from the summer when youth are more likely to be outdoors in brighter light environments, to shift into darker indoor artificial lighting environments.

Limitations of this work should be considered. Since this is a cross-sectional study, we cannot determine whether these findings are a result of patients with headache-related photophobia being more likely to present in the winter, or whether symptoms of photophobia are worse in the winter at the individual level. Longitudinal studies are needed to address this question.

Identifying the presence or absence of photophobia was based on patient report, which likely under-estimate rates of photophobia, as suggested by our prior work using this headache registry.^14^ However, this is also a strength of this work, as leveraging patient-report may have allowed us to uncover seasonal differences, with those who experience photophobia being less likely to report these symptoms when they are less bothersome. Additionally, it is unclear if it is the overall intensity of light exposure, or if time indoors in artificial lighting environments evokes more visual discomfort, which would require real-world measurements of daily light exposure. Further study is needed to understand the relationship between increased rates of photophobia in the winter and determine the generalizability of this result in different geographic locations and adult populations.

## Data Availability

All data produced in the present study are available upon reasonable request to the authors.

## Abbreviations

CHOP: Children’s Hospital of Philadelphia
ICHD-3: International Classification of Headache Disorders 3^rd^ Edition
IQR: Interquartile range

## Acknowledgements

We would like to thank the patients of the Children’s Hospital of Philadelphia General Neurology and Headache Clinics for taking the time to fill out the questionnaire.

